# DeltaScan: 1-minute single-channel EEG, a valuable tool for diagnosing delirium in frail older patients in the emergency department

**DOI:** 10.1101/2025.08.01.25332717

**Authors:** Loes Caelers, Steffie Peeters, Laurien Theunissen, Guy Mostard, Mark Klein Ovink, Maurice Janssen, Ellen Castro, Chantal de Weerd Spaetgens, Dennis Kurstjens, Audrey Merry, Walther Sipers

## Abstract

**Introduction:** Diagnosing delirium is challenging, especially in the emergency department (ED), where up to 80% of diagnoses are missed. The DeltaScan is a portable EEG-based delirium monitor that makes a single-channel EEG recording and quantifies delirium on a 5-point scale. The aim of this study was to determine whether the DeltaScan is an accurate diagnostic instrument for delirium in frail, older patients that present at the ED.

**Method and materials:** For this cross-sectional study, data were collected between July 2022 and December 2022 at the ED of a non-academic teaching hospital in the Netherlands. Patients were included if they were older than 70 and frail. The included patients underwent the DeltaScan measurement. The results of the DeltaScan were compared with the results of the Confusion Assessment Method assessed by delirium experts.

**Results:** A total of 104 patients were included in this study. The mean age was 84.9 years and 55.4% were female. Because of technical difficulties 3 patients had to be excluded. There was no diagnostic consensus between delirium experts in 21 patients. In the remaining 80 patients, delirium was diagnosed in 41%. The sensitivity of the DeltaScan was 81.3% (67.7-94.8) and the specificity was 89.6% (80.9-98.2). The positive predictive value was 83.9% (70.9-96.8) and the negative predictive value was 87.8% (78.6-96.9). The area under the receiver operating characteristics curve was 86.5% (77.5-96.6).

**Conclusion:** Diagnosing a delirium in frail, older patients attending the ED can be challenging, even for delirium experts. We demonstrated that DeltaScan measurement is an objective, accurate and feasible approach to diagnose delirium. We therefore argue that the DeltaScan should have a permanent place for diagnosing delirium in the ED.

## Introduction

Diagnosing delirium is challenging, especially in the emergency department (ED), where up to 80% of diagnoses are being missed [1]. One explanation for these missed diagnoses is that diagnostic instruments are subjectively interpreted by means of the clinical observation and the interpretation is therefore highly dependent on the clinical experience of the treating physicians. Physicians that work in the ED are often relatively young with little experience in recognizing delirium and differentiating delirium from dementia and other cognitive disorders. Furthermore, using these diagnostic instruments is time-consuming. Performing the Confusion Assessment Method (CAM) for example, which is the most well studied clinical delirium assessment tool, takes 7 to 15 minutes. *However, even experienced physicians often disagree about the presence or absence of delirium and therefore a high interobserver variability is seen [2].* In the ED, the focus is on acute illness and due to the high turnover there is often limited time to do a full cognitive geriatric assessment. Nevertheless, especially in the ED it is of vital importance that the diagnosis is accurate. Delirium is a signal of an underlying condition. These conditions often present atypically in older people in which delirium may be the only symptom present. Among patients with delirium, the overall mortality rate is increased threefold and is comparable to the mortality rate in patients with myocardial infarction or sepsis [3]. Delay in starting treatment worsens the outcome [4,5].

An objective approach to detect delirium is to monitor physiological changes. During delirium, electroencephalography (EEG) shows a slowdown in background activity, the ‘polymorphic delta waves’ [6,7,8]. Performing a conventional 25-electrode EEG in all patients at risk for delirium is unsuitable for daily clinical practice because it is burdensome, requires trained personnel for interpretation and is therefore expensive. Previous research showed that delirium could be adequately assessed using a single channel EEG looking at delta waves [9]. A proof-of-concept study with 28 postoperative cardiothoracic patients showed that the relative deltapower of a one-minute EEG recording with only two electrodes can distinguish between patients who have delirium and those who do not [10]. Based on these findings, Ditzel et al. developed and validated the DeltaScan algorithm in a study with 145 patients who underwent major surgery [11]. This study showed a diagnostic performance of >85% (AUC=0.86, CI = [0.81–0.90]). To improve the feasibility of the measurement of the DeltaScan algorithm, the DeltaScan was developed (Prolira^®^ Arnhem, the Netherlands). The DeltaScan is a portable EEG-based delirium monitor that makes a single-channel EEG recording in 1 minute. Based on this EEG recording the DeltaScan quantifies delirium on a 5-point scale.

However, the diagnostic performance of the DeltaScan has not yet been investigated in patients in the ED. Furthermore, the DeltaScan has not been validated in patients with cognitive disorders such as dementia. Patients presenting at the ED with a possible delirium are a heterogeneous group with various acute illnesses and comorbidities such as dementia are frequent. They are generally older, frail and polypharmacy is common in this population.

The aim of this study was to determine whether the DeltaScan is an accurate diagnostic instrument for delirium in frail, older patients with and without cognitive disorders that present at the ED. Furthermore, we aimed to determine whether the DeltaScan is time efficient and feasible in patients with possible cognitive disorders and agitation that might complicate the DeltaScan measurement. We hypothesize that the DeltaScan is a feasible, fast, and accurate diagnostic instrument not only in well-defined study cohorts but also in a heterogeneous, older, and frail population.

## Methods

### Study setting and population

This cross-sectional study was done in a non-academic teaching hospital in the Netherlands. The data were collected at the ED between July 2022 and December 2022. Patients were included if they were older than 70 years, frail according to the Groningen Frailty Indicator (GFI, frail if score ≥4) or according to the Identification of the Seniors At Risk (ISAR, frail if score ≥2) and if they were referred to the ED for the specialties geriatric medicine, internal medicine or emergency medicine. Patients were excluded if they were terminally ill, if it was impossible to take cognitive tests due to deafness or a language barrier, if head bandage prevented the placement of electrodes, if they used lithium or clozapine, if macro brain injury during the 6 weeks before referral to the ED had occurred or if they had a metal plate or device in cerebro. Patients were also excluded if severe agitation interfered with the DeltaScan measurement. Informed consent was obtained from all participants or their legal representatives.

### Data collection and classification

The two researchers, SP and LT, which are medical students in the final master phase, included patients at the ED that met the inclusion criteria. Of the included patients, patient characteristics were recorded on the patient report file (PRF), including referral information, patient history, anamnesis, hetero anamnesis, and additional research that was done in the ED like laboratory diagnostics, electrocardiogram and imaging.

For this study we used the Prolira^®^ (Arnhem, The Netherlands) DeltaScan Brain State Monitor (serial number: 20050009, algorithm version 2.4.2, DeltScan Patch version: REF 009.000.B, Lavel V2B). The DeltaScan is an EEG-based delirium monitor that makes a single-channel EEG recording in one minute using three electrodes. The DeltaScan quantifies a delirium on a 5-point scale. Score 1 means very unlikely delirium, 2 unlikely delirium, 3 possible delirium, 4 likely delirium and 5 means very likely delirium. We categorized a DeltaScan score of 1 or 2 as no delirium and a score of 3, 4, or 5 as delirium.

The two researchers were trained by an expert researcher affiliated with Prolira^®^ to use the DeltaScan equipment. The expert supervised the first measurements and was available for troubleshooting in case of device-related questions or problems. The researchers visited the included patients at the ED to perform a cognitive assessment based on the CAM, which was recorded on video. Immediately after the cognitive assessment, the DeltaScan measurement was performed. Since noise, such as eye movements and glossokinetic artifacts predominantly manifests within the low-frequency range of the DeltaScan, the researcher ensured that during the 1-minute recording the patients had their eyes closed and did not speak.

The videos were assessed by the delirium experts if the score of the DeltaScan was 2, 3 or 4, or if there was no consensus about the presence or absence of delirium between the ED resident, the attending geriatrician, and the researcher. If the score of the DeltaScan was 1 or 5, and there was consensus between the ED resident, the attending geriatrician and the researcher, the diagnosis was confirmed, and no further video assessment was done. The delirium experts were two independent geriatricians with at least 8 years of experience. They were blinded from the DeltaScan score and the conclusion of the researcher, ED resident and attending geriatrician. After examining the PRF and the videos they classified diagnosis based on the CAM criteria (gold standard) in the following five options: 1) clinical picture of a delirium with features of a delirium at that moment, with actual at least 3 positive CAM criteria, 2) clinical picture of a delirium with some delirious features at that moment, with at least 1 actual CAM criterium, 3) clinical picture of a delirium with doubtful delirious features at that moment, without an actual CAM criterium, however with clinical information high suspicious for a delirium, 4) possible delirium, with clinical information with some hallmarks of a delirium 5) no delirium. We categorized a score of 1-4 as delirium and a score of 5 as no delirium according the CAM criteria. If there was an agreement between the two delirium experts, the conclusion was added to the results. Patients were not included in the analysis if there was no agreement between the delirium experts due to uncertainty about the diagnosis.

### Outcome measures

The primary outcome of our study was the diagnostic value of the DeltaScan in terms of sensitivity, specificity, positive predictive value (PPV), negative predicting value (NPV), receiver operating characteristic curve (ROC curve) and area under the ROC Curve (AUC), using the diagnosis made by the independent delirium experts based on the CAM criteria as ‘gold standard’. Furthermore, we wanted to determine the feasibility of the DeltaScan by measuring the number of patients that were excluded due to technical difficulties and severe agitation that interfered with the DeltaScan measurement.

### Sample size calculation

We did not perform a power analysis in advance because due to financial reasons there were 110 electrode-patches available.

### Statistical analysis

Analysis was performed in SPSS (SPSS Statistics, version 26). Categorical variables were presented as numbers with the corresponding percentages, continuous variables were presented as mean with Standard Deviation (SD) (normal distribution).

Differences between the patients with a positive DeltaScan score and a negative DeltaScan score were determined by means of the Chi-square test and the Fisher-Freeman-Halton exact test for categorical variables. The independent sample-t-test was applied to compare continuous variables with a normal distribution. If not normally distributed, the Mann-Whitney U test was used. Receiver operating characteristics (ROC) curve was computed and the area under the ROC curve (AUC) was calculated with accompanying 95% confidence interval (CI). In addition, we calculated the PPV, NPV, sensitivity, and specificity for the DeltaScan scores/diagnoses using the diagnoses made by the delirium experts as reference. Feasibility was determined by evaluating the number of patients excluded due to technical failure of the DeltaScan and due to severe agitation that interfered with the DeltaScan measurement. Also, the time it took to learn how to use the DeltaScan and the time consumption of the DeltaScan measurement were considered.

The study was approved by the Medical Ethics Committee of Zuyderland and Zuyd University of Applied Sciences (protocol METCZ20220008).

## Results

### Study population

Figure 1 shows the trial profile of the research population. Initially, 104 patients met the inclusion criteria. Due to technical difficulties with the DeltaScan, 3 patients were excluded. No patients were excluded due to severe agitation that interfered with the DeltaScan measurement. Of the 101 patients that were initially assessed, the DeltaScan score was 1 or 5 in 65 cases (64.4%). In 47 of these cases, there was consensus about the diagnosis between the ED resident, the attending geriatrician and the researcher and the videos were not assessed by the delirium experts. In the remaining 18 cases in which there was no consensus about the diagnosis, and in the 36 cases in which the DeltaScan score was 2, 3 or 4, the videos were assessed by the delirium experts. In 33 of these 54 cases, there was consensus between the 2 delirium experts about the presence or absence of delirium. This resulted in a definite conclusion in 80 patients.

**Figure 1.**
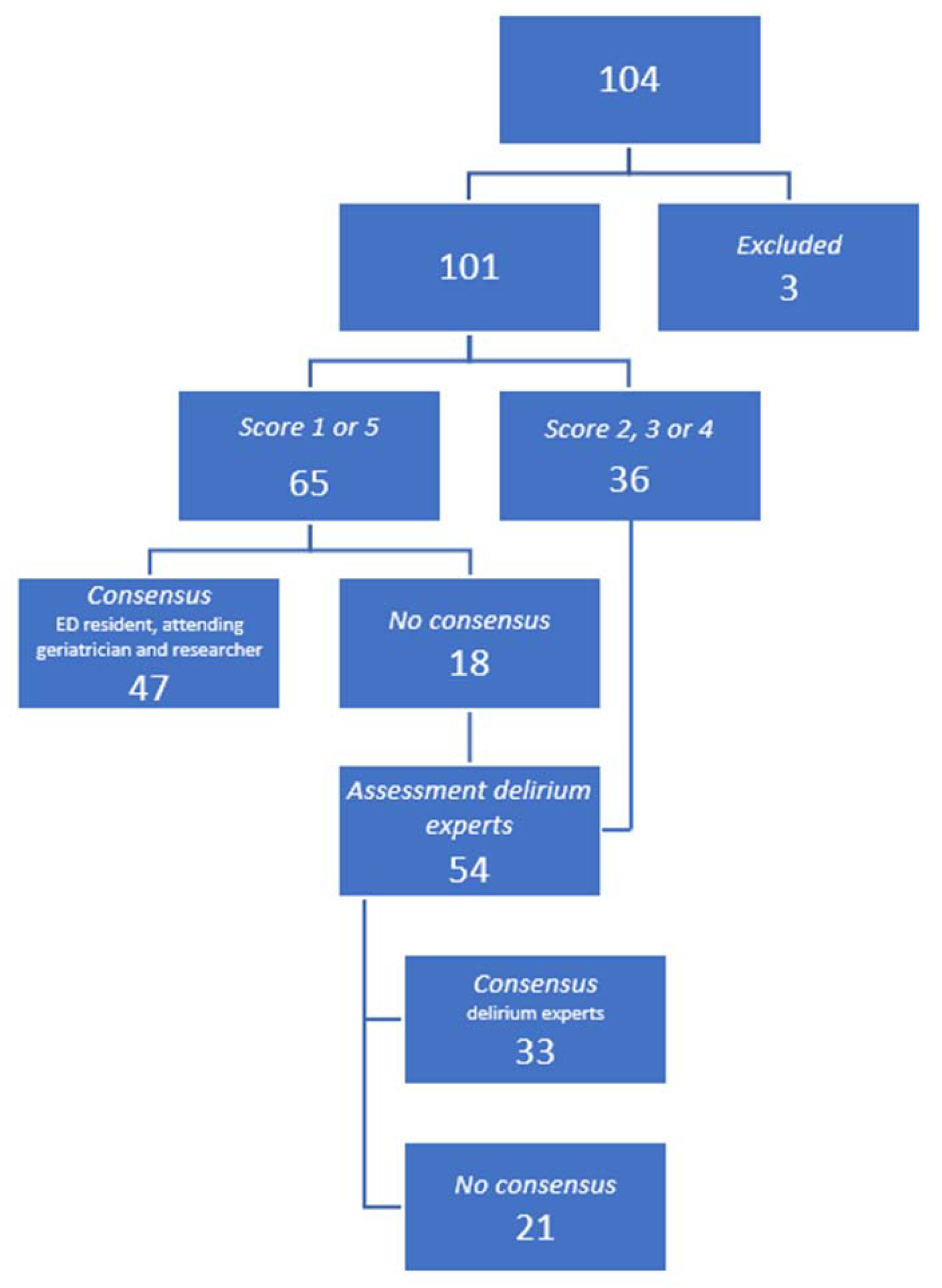
Flowchart of inclusion of frail older patients at the ER in diagnosing delirium. Score 1-5 means CAM score: 1) clinical picture of a delirium with features of a delirium at that moment, with actual at least 3 positive CAM criteria, 2) clinical picture of a delirium with some delirious features at that moment, with at least 1 actual CAM criterium, 3) clinical picture of a delirium with doubtful delirious features at that moment, without an actual CAM criterium, however with clinical information high suspicious for a delirium, 4) possible delirium, with clinical information with some hallmarks of a delirium 5) no delirium.

### Baseline characteristics

Supplementary Table 1 shows the baseline characteristics of the included patients (n=101) and the patients with consensus about the diagnosis of delirium according to reference (n=80). The mean age was 84.9 years and 55.4% were female of the included patients. A history of cerebrovascular accidents was found in 29.7% and a history of delirium was found in 16.8% of the patients. Dementia occurred in 22.8% of the patients. Psychopharmaceutic drugs were used by 35.6% of the patients; in 9.9% benzodiazepines were used. These characteristics did not differ significantly for the patients with (n=80) and without diagnostic consensus (n=21). Also, there were no significant differences between the patients with and without delirium according to the DeltaScan (*Table 1*).

**Table 1.**
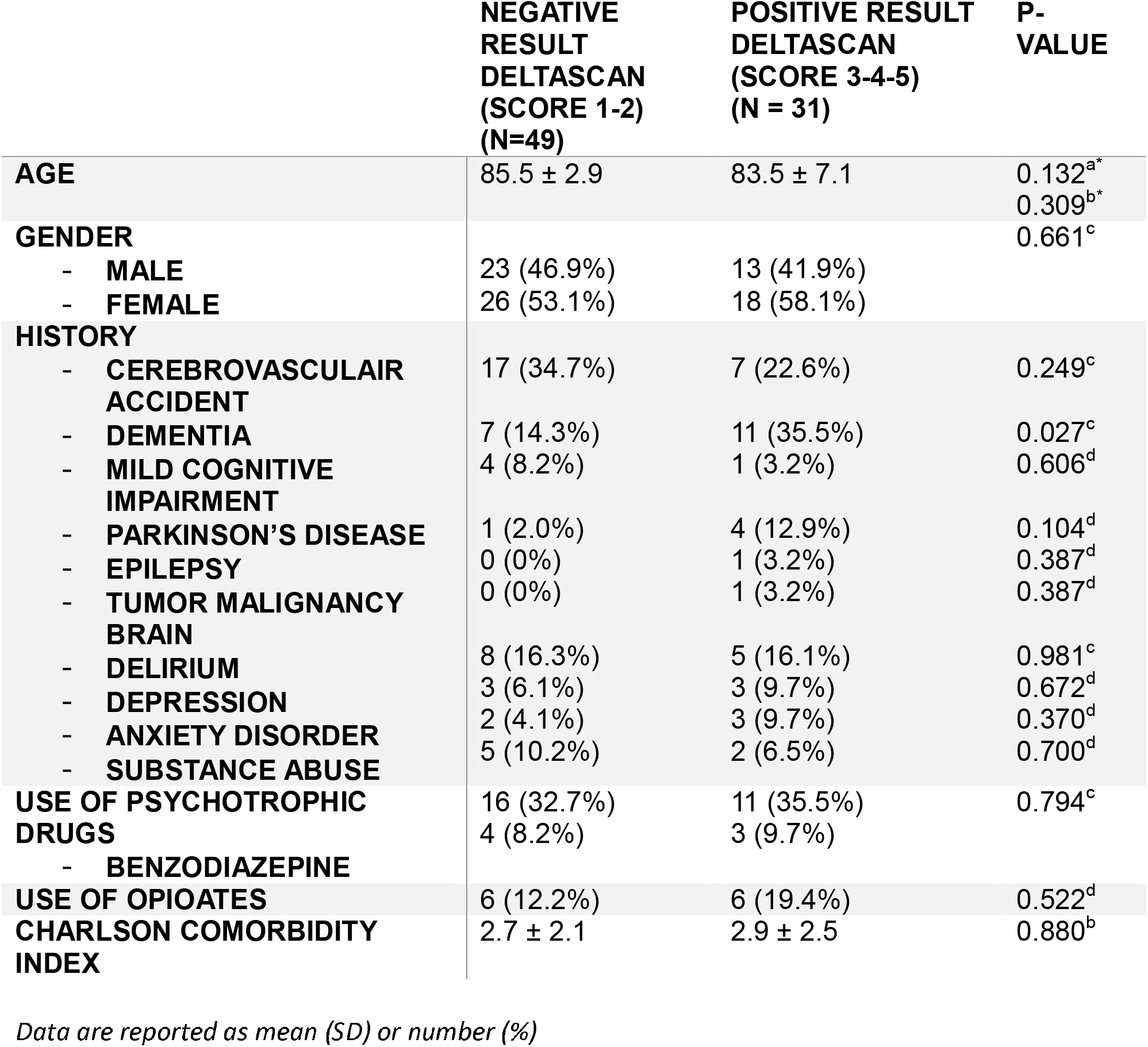
Comparison of baseline characteristics of the patients with a positive (n=31) and negative result (n=49) on the DeltaScan (n=80)

### Outcome

Table 2 shows an overview of the results of the DeltaScan compared to the reference. There were 26 patients with a positive score on the DeltaScan (score 3,4 or 5) and a positive diagnosis according to the CAM criteria, 5 patients with a positive result on the DeltaScan and a negative diagnosis according to the CAM criteria, 7 patients with a negative result on the DeltaScan (score 1 or 2) and a positive diagnosis according to the CAM criteria and 42 patients with a negative result on the DeltaScan and a negative diagnosis according to the CAM criteria. Sensitivity was 81.3% (CI 67.7%-94.8%) and specificity was 89.6% (CI 80.9%-98.2%). PPV was 83.9% (CI 70.9%-96.8%) and NPV was 87.8% (CI 78.6%-96.9%). Figure 2 shows the ROC curve of the diagnostic value of the DeltaScan compared to the reference. The area under the ROC curve (AUC) is 86.5% (CI 77.5%-95.6%).

**Table 2.**
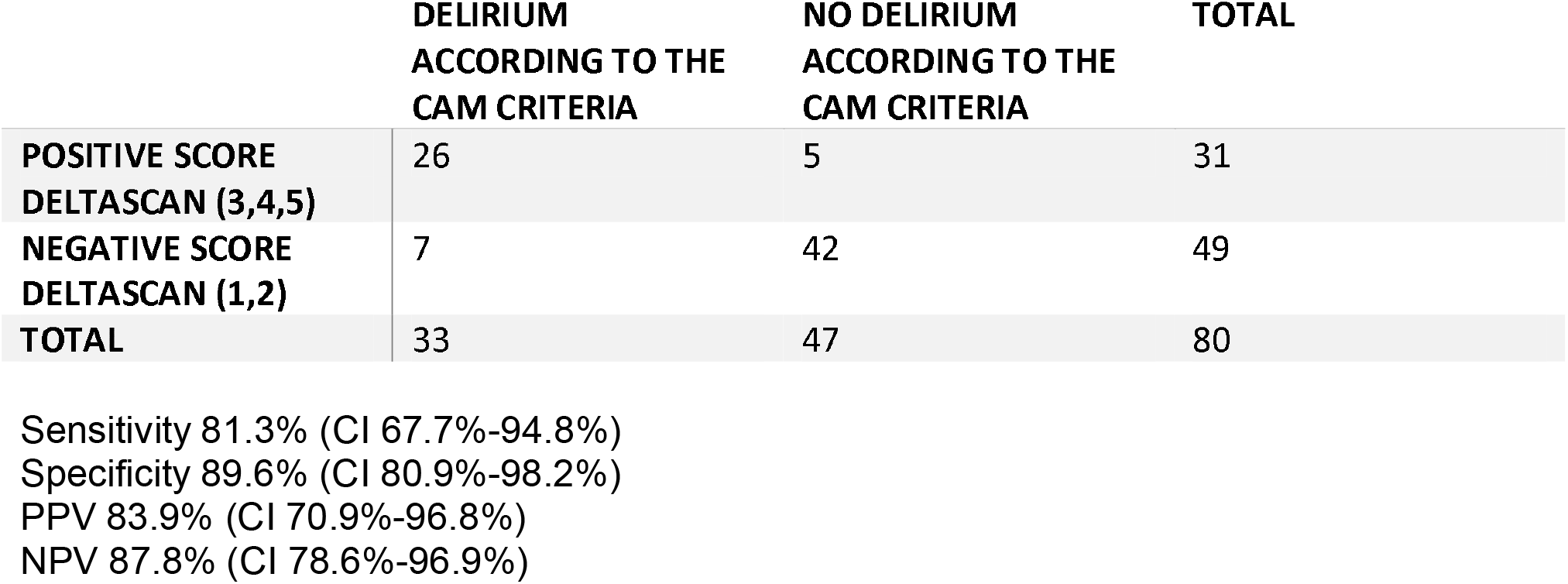
Results of DeltaScan score compared with diagnosis delirium according with CAM criteria with consensus reference in frail older patients (n=80) in the ED.

|  | **DELIRIUM**<br>**ACCORDING TO THE**<br>**CAM CRITERIA** | **NO DELIRIUM**<br>**ACCORDING TO THE**<br>**CAM CRITERIA** | **TOTAL** |
| --- | --- | --- | --- |
| **POSITIVE SCORE**<br>**DELTASCAN (3,4,5)** | 26 | 5 | 31 |
| **NEGATIVE SCORE**<br>**DELTASCAN (1,2)** | 7 | 42 | 49 |
| **TOTAL** | 33 | 47 | 80 |

Sensitivity 81.3% (CI 67.7%-94.8%)
Specificity 89.6% (CI 80.9%-98.2%)
PPV 83.9% (CI 70.9%-96.8%)
NPV 87.8% (CI 78.6%-96.9%)

**Figure 2.**
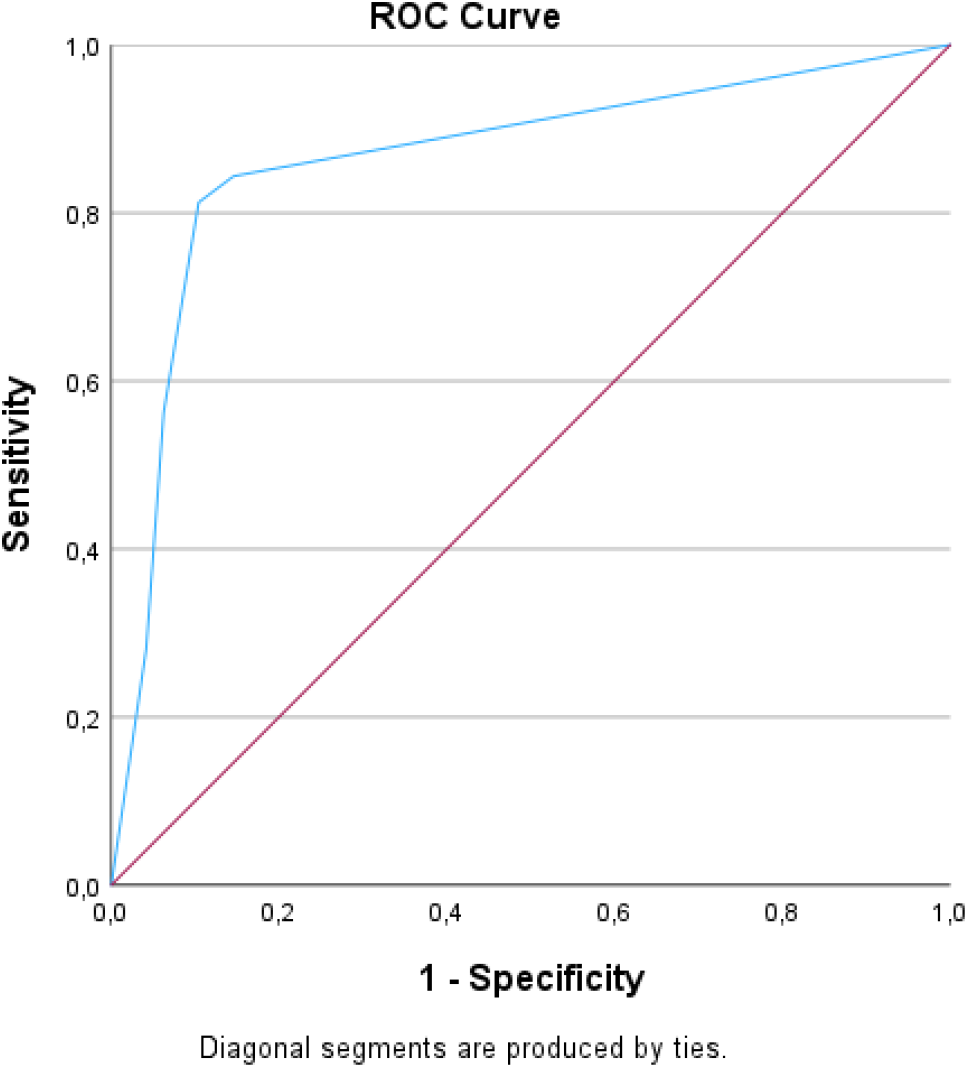
ROC curve of the DeltaScan in diagnosing delirium according CAM criteria in frail older patients (n=80) in the ED. AUC of 86.5% (95% CI 77.5-95.6)

Table 3 presents an overview of the individual DeltaScan scores versus the individual classifications according to the assessments of the delirium experts. In case of a delirium diagnosis according to the reference in the presence of delirium features on the moment of measurement, DeltaScan score is positive in 21 of 23 patients (91%). In case of a delirium diagnosis according to the reference in absence of delirium features on the moment of measurement, DeltaScan score is positive in 1 of 4 patients (25%).

**Table 3.** Overview of frail older patients (n=80) in the ED with individual DeltaScan scores versus individual groups reference diagnosis according CAM criteria.

| SCORE<br>DELTASCAN | RESULT REFERENCE ACORDING CAM CRITERIA |  |  |  |  | TOTAL<br>PATIENTS |
| --- | --- | --- | --- | --- | --- | --- |
|  | No<br>delirium<sup>5</sup> | Delirium +<br>features<sup>1</sup> | Delirium –<br>features<sup>2</sup> | Delirium<br>+/-<br>features<sup>3</sup> | Doubtful<br>delirium<sup>4</sup> |  |
| 1 | 41 | 2 | 3 | 0 | 0 | 46 |
| 2 | 1 | 0 | 0 | 1 | 1 | 3 |
| 3 | 2 | 5 | 1 | 1 | 1 | 10 |
| 4 | 1 | 7 | 0 | 1 | 1 | 10 |
| 5 | 2 | 9 | 0 | 0 | 0 | 11 |
| **TOTAL**<br>**PATIENTS** | 47 | 23 | 4 | 3 | 3 | 80 |

<sup>5</sup> No delirium diagnosis, also no features of delirium present. <sup>1</sup> Delirium diagnosis with features of a delirium at that moment, with actual at least 3 positive CAM criteria. <sup>2</sup> Delirium diagnosis with some delirious features at that moment, with at least 1 actual CAM criterium. <sup>3</sup> Delirium diagnosis with doubtful delirious features at that moment, without an actual CAM criterium, however with clinical information high suspicious for a delirium. <sup>4</sup> Doubtful delirium diagnosis, with clinical information with some hallmarks of a delirium

The time it took to learn how to use the DeltaScan was less than 1 hour. The time consumption of the DeltaScan measurement was 5 minutes. None of the patients had to be excluded due to severe agitation that complicated the DeltaScan measurement.

## Discussion

The aim of this study was to determine whether the DeltaScan is a feasible and accurate diagnostic instrument for delirium in frail, older patients with and without cognitive disorders attending the ED. Diagnosing delirium can be challenging and therefore diagnoses are often missed.

We included 101 patients (mean age 84.9 years) from which 22.8% had dementia. The results of the DeltaScan were compared with the results of the Confusion Assessment Method based on consensus between researchers and attending physician or delirium experts. Our study demonstrates that the DeltaScan is a feasible and accurate instrument for diagnosing delirium in frail, older patients at the ED. Even though 3% (3/104) of the patients had to be excluded due to technical difficulties of the DeltaScan, none of the patients had to be excluded due to severe agitation that complicated the DeltaScan measurement. Also, performing the DeltaScan measurement took only 5 minutes. This shows that the DeltaScan is a feasible instrument, even when patients have cognitive disorders in which case they might not be able to understand instructions. The technical difficulties occurred early in the study and were caused by an outdated version of the electrode-patches. During the study, the electrodes were replaced by a new series and no further technical problems occurred. Furthermore, our study demonstrates that the DeltaScan is an accurate diagnostic instrument, with a sensitivity of 81.3% (CI 67.7%-94.8%) and a specificity of 89.6% (CI 80.9%-98.2%). The PPV was 83.9% (CI 70.9%-96.8%) and the NPV was 87.8% (CI 78.6%-96.9%). The AUC was 86.5% (CI 77.5-95.6).

Our findings confirm the assumptions of previous studies, which found comparable diagnostic values of the DeltaScan in well-selected patients and confined study environments [9,11,12]. There are several differences between previous studies and the present study. Most importantly, previous studies were done in well-defined study cohorts while the present study was done in a heterogenous, frail, older population in the ED. In contrast to our study, in previous research populations patients with cognitive disorders or dementia were either excluded [10,13] or not likely prevalent due to relatively young research populations that had underwent major surgery or were admitted to the ICU [2,9,11,12,14], both interventions that are generally too invasive in frail patients with cognitive disorders or dementia. There are several distinct diseases that are classified under the umbrella term ‘dementia’, and they have different, specific electroencephalogram characteristics. Overall, typical findings in the various types of dementia are an increase in EEG slow wave activity, e.g. theta and delta frequencies, together with decreased activity in the alpha frequency [14]. The study was not primarily designed to investigate the diagnostic performance of the DeltaScan in patients with dementia, therefore we cannot make any further statements about this. We simply choose not to exclude patients with a dementia to make our results more generalizable to daily practice because of the high prevalence of cognitive disorders and dementia in frail, older patients that present at the ED [15,16]. Moreover, especially in case of cognitive disorders, diagnosing delirium can be challenging due to overlap in symptoms and clinical presentation, which emphasizes the need for a better diagnostic instrument in this patient group. Also, in many cases, dementia is not recognized and diagnosed late [17], which means that a possible diagnosis of dementia is not always known at the time of presentation at the ED and therefore exclusion is not always possible.

Diagnosing delirium in the ED by using the DeltaScan is objective and time efficient. Moreover, specific expertise in recognizing and diagnosing delirium is not required. In our study, the two researchers were trained both theoretically and practically by an expert researcher affiliated with Prolira^®^ to use the DeltaScan equipment, which took only 60 minutes in total. After that, they were able to perform the entire measurement on their own in 5 minutes. Furthermore, no background knowledge about the EEG and its interpretation is required because the DeltaScan contains an independently functioning algorithm. Our findings suggest that this is an approach that could be used to diagnose delirium by bedside nurses at the ED as part of the clinical routine, especially at the ED setting where nurses already monitor a broad range of information. Since delirium is frequently missed in the ED, the DeltaScan can make an important contribution to the improvement of the quality of care for frail, older patients in the ED.

Clearly, there are limitations of the present study. One of the main difficulties when investigating a diagnostic instrument for delirium is the lack of a flawless gold standard. Studies on delirium detection tools usually have a delirium expert or a panel of delirium experts as reference. However, experts frequently disagree about the diagnosis of delirium, even when considering the same clinical and cognitive information [2]. Even though delirium experts involved in our study had extensive experience in diagnosing delirium, there was no consensus when applying the CAM criteria between the experts in 21% of the cases. In previous research this number was lower (Numan 2018). In our opinion the lack of a perfect gold standard and the level of disagreement between delirium experts only emphasize the need for an objective instrument like the DeltaScan. To overcome this problem in patients with altered behavior and a low score on the DeltaScan, one or more follow-up measurements should be considered to be certain whether this low score is caused by a fluctuation in the condition and to be certain of a correct diagnosis. However, this should be investigated further.

Furthermore, EEG recordings were performed only once, which might have resulted in overlooking periods of slowing of the EEG. One of the key features of delirium is its fluctuating course. It is a possibility that the DeltaScan score is influenced by the presence or absence of delirium features at the time of the measurement. This might explain that, as can be seen in table 3, in case of delirium according to the reference, DeltaScan score is positive in 21/23 (91.3%) patients with delirium with at least 3 CAM criteria, in contrast to 1/4 (25%) patients with delirium without delirium features at that moment. Therefore, in patients with persistent behavioral changes, a repeat of the DeltaScan measurement seems appropriate. For clinical practice, as suggested above, a diagnostic algorithm in applying the DeltaScan could be of value.

Finally, it could be argued that the observed differences in EEG characteristics between patients classified with or without delirium were influenced by differences in drowsiness, sleep or administration of benzodiazepines or opioids. Furthermore, noise, such as eye movements and glossokinetic artifacts, predominantly manifests within the low-frequency range of the EEG and can therefore influence the DeltaScan measurement. However, use of psychotropic drugs, benzodiazepines and opiates did not differ between the patients with a positive DeltaScan score and the patients with a negative DeltaScan score. In addition, polymorphic delta activity caused by sleep or noise seems unlikely since the researcher constantly ensured that the patients were awake, had their eyes closed and did not speak during the measurement.

## Conclusion

Diagnosing a delirium in frail, older patients that present at the ED can be challenging, even for delirium experts. The present study demonstrated that DeltaScan measurement is an objective, accurate and feasible approach to diagnose delirium in these patients in the ED. We therefore argue that the DeltaScan should have a permanent place for diagnosing delirium in the ED.

## Data Availability

All relevant data are within the manuscript and its Supporting Information files.

## Acknowledgments

In addition to the authors, we thank Erik Tio, clinical specialist DeltaScan from Prolira for training our researchers and his availability for troubleshooting in case of device-related questions or problems

## References

1. Lewis LM, Miller DK, Morley JE, Nork MJ, Lasater LC. Unrecognized delirium in ED geriatric patients. Am J Emerg Med 1995; 13: 142–5.

2. Numan T, van den Boogaard M, Kamper AM et al. Recognition of Delirium in Postoperative Elderly Patients: A Multicenter Study. J Am Geriatr Soc 2017; 65: 1932–1938.

3. Ely EW, Shintani A, Truman B et al. Delirium as a predictor of mortality in mechanically ventilated patients in the intensive care unit. JAMA 2004; 291: 1753–62.

4. Marcantonio ER. Delirium in Hospitalized Older Adults. N Engl J Med 2017; 377: 1456–1466.

5. Slooter AJC, Van De Leur RR, Zaal IJ. Delirium in critically ill patients. Handb Clin Neurol 2017; 141: 449–466.

6. Koponen H, Partanen J, Pääkkönen A, Mattila E, Riekkinen PJ. EEG spectral analysis in delirium. J Neurol Neurosurg Psychiatry 1989; 52: 980–5.

7. Hut SCA, Dijkstra-Kersten SMA, Numan T et al. EEG and clinical assessment in delirium and acute encephalopathy. Psychiatry Clin Neurosci 2021; 75: 265–266.

8. Pandharipande PP, Ely EW, Arora RC et al. The intensive care delirium research agenda: a multinational, interprofessional perspective. Intensive Care Medicine 43 2017 1329–1339.

9. Numan T, van den Boogaard M, Kamper AM et al. Delirium detection using relative delta power based on 1-minute single-channel EEG: a multicentre study. Br J Anaesth 2019; 122: 60–68.

10. van der Kooi AW, Zaal IJ, Klijn FA et al. Delirium detection using EEG: what and how to measure. Chest 2015; 147: 94–101.

11. Ditzel FL, Hut SC, Dijkstra-Kersten SM et al. An automated electroencephalography algorithm to detect polymorphic delta activity in acute encephalopathy presenting as postoperative delirium. Psychiatry Clin Neurosci 2022; 76: 676–678.

12. Aben J, Pouwels S, Oldenbeuving A. Comparison Between Deltascan Single Channel Electroencephalography (EEG), Confusion Assessment Method-Intensive Care Unit (CAM-ICU) Score and Clinical Assessment in Diagnosing Delirium in Intubated Patients in the Intensive Care Unit. Cureus 2022; 14: e26449.

13. Hut SCA, Dijkstra-Kersten SMA, Numan T et al. EEG and clinical assessment in delirium and acute encephalopathy. Psychiatry Clin Neurosci 2021; 75: 265–266.

14. Malek N, Baker MR, Mann C, Greene J. Electroencephalographic markers in dementia. Acta Neurologica Scandinavica 135 2017 388–393.

15. Hirschman KB, Paik HH, Pines JM, McCusker CM, Naylor MD, Hollander JE. Cognitive Impairment among Older Adults in the Emergency Department. West J Emerg Med 2011; 12: 56–62.

16. LaMantia MA, Stump TE, Messina FC, Miller DK, Callahan CM. Emergency Department Use Among Older Adults With Dementia. Alzheimer Dis Assoc Disord 2016; 30: 35–40.

17. Amjad H, Roth DL, Sheehan OC, Lyketsos CG, Wolff JL, Samus QM. Underdiagnosis of Dementia: an Observational Study of Patterns in Diagnosis and Awareness in US Older Adults. J Gen Intern Med 2018; 33: 1131–1138.

